# Predicting the Association Between Oral Hyperpigmentation and Pregnancy in Women Aged 18-45 Using Machine Learning and Directed Acyclic Graphs: A Cross-Sectional Study

**DOI:** 10.64898/2026.01.06.26343400

**Authors:** Hifza Noor, Arooba Malik, Mahnoor Fatima, Rameesa Aymen, Faisal Shafiq Malik, Momina Zahra

**Affiliations:** Riphah International University, Islamabad, Pakistan; Assistant Professor, Community Dentistry, Riphah International University, Islamabad, Pakistan

**Keywords:** Algorithms, Cross-Sectional Studies, Maternal Health, Regression Analysis, Oral Health

## Abstract

**Objectives:** The study aims to predict the association between oral hyperpigmentation and pregnancy in women between 18 and 45 years using machine learning (ML) algorithms. Directed acyclic graphs (DAGs) were constructed to explore the association between oral hyperpigmentation and demographic factors such as age, occupation, ethnicity, and number of pregnancies.

**Methods:** This cross-sectional study involved pregnant and non-pregnant participants aged between 18 and 45. The study was conducted at one public and one private sector hospital. DAGs were used to represent relationships between different variables. Three ML models, including Logistic Regression (LR), Random Forest (RF) and Gradient Boosting Machine (GBM) algorithms, were used to predict the association between oral hyperpigmentation and pregnancy based upon relevant theoretically backed predictors.

**Results:** The results show that labial mucosa and hard palate hyperpigmentation had a positive correlation with the number of pregnancies, with p-values of 0.003 and 0.018, respectively. All three ML models demonstrated excellent predictive power. However, the LR model with an accuracy rate of 96.67% was particularly effective in predicting the association between oral hyperpigmentation and pregnancy.

**Conclusions:** This study highlighted the effectiveness of LR, supported by DAGs, in predicting oral hyperpigmentation among women aged 18–45. Key factors such as the number of pregnancies, age, ethnicity, and occupation significantly influenced site-specific hyperpigmentation. The findings support the integration of ML into maternal oral healthcare for early detection and culturally tailored interventions. Simpler ML models proved most accurate in moderate-sized datasets with fewer covariates.

**Significance:** Oral hyperpigmentation is known to be impacted by pregnancy, though few predictive models incorporate demographic factors into consideration. This study uses DAG-supported machine learning models to investigate and predict the association between oral hyperpigmentation and pregnancy in women of age 18 to 45 years. This study contributes to data-driven maternal healthcare by developing predictive models tailored to the ethnically diverse population, aiming to improve patient counselling, early detection, and guide preventive measures in oral healthcare for females.

## Introduction

Oral hyperpigmentation is a physiological phenomenon due to the excess deposition of melanin and other pigments on the oral mucosal surfaces (Abati, 2024). It is mostly undetectable and presents as discoloration of the gingiva, buccal mucosa, or other intraoral sites (Agarwal et al., 2015). This hyperpigmentation, due to its resemblance with pathological lesions and aesthetic concerns, especially for pregnant women facing hormonal changes, serves as a diagnostic challenge (Abati, 2024; Cicek & Ertas, 2003; Pecci-Lloret et al., 2024).

The prevalence of oral hyperpigmentation changes considerably amongst the population, occurring in approximately 20.8% of people worldwide (95% CI: 17.1–25.0%) (Rotbeh et al., 2022). Besides genetic, environmental, social, and cultural factors influencing oral hyperpigmentation, this condition is influenced by physiological factors like age, occupation, ethnicity, and the number of pregnancies (Gulati et al., n.d.). For example, people with darker skin tones, such as those of Africans and South Africans, tend to have more oral hyperpigmentation than people with lighter skin tones (Masilana et al., 2017; Ponnaiyan et al., 2024; Feller et al., 2014). A study carried out in South Africa, with a prevalence of 42.3% and 430 subjects from various ethnic backgrounds, reported a greater detection rate for orofacial hyperpigmentation (Masilana et al., 2017). Similarly, oral hyperpigmentation patterns in developing countries are strongly affected by genes due to the diverse population in terms of race and ethnicity (Almazrooa et al., 2025).

Environmental exposures, including smoking, sun exposure, and air quality, play an important part in hyperpigmentation. It is known that ultraviolet (UV) radiation affects the synthesis of melanin (Abati, 2024). In areas of developing countries that get a lot of UV radiation, people typically experience greater levels of skin and oral hyperpigmentation (Shahzad et al., 2018). Similarly, brown to black discoloration of the oral mucosa in smoker’s melanosis, particularly in the buccal, hard palate, and gingiva, has a link with smoking (Abati, 2024). A precise diagnosis and patient reassurance, specifically for populations at risk, require an understanding of its patterns and causes.

Hormonal changes during pregnancy, notably high levels of progesterone and estrogen, promote melanin to be produced in excess, which results in hyperpigmentation in different parts of the body (Pecci-Lloret et al., 2024). The areas involved included the face (*melasma*) and abdomen (*Linea nigra*) (Motosko et al., 2017). However, studies on the association of pregnancy and oral hyperpigmentation are extremely limited in developing countries, with no integrated models developed to predict these changes. This represents a major gap in women’s oral health research (Sattar et al., 2017).

While studying oral hyperpigmentation, it was observed that the traditional analysis methods frequently lack the ability to detect the complex nature of these interlinked environmental, physiological, and demographic factors. Oral hyperpigmentation research still underutilizes advanced techniques like Directed Acyclic Graphs (DAGs) and Machine Learning (ML), in spite of their increasing relevance in the healthcare domain (Dubuc et al., 2022). While DAGs enable the clear visualization of causal interactions and enable researchers to differentiate between confounders, mediators, and colliders and avoid statistical bias, ML algorithms are capable of detecting non-linear associations and generating predictive insights. Together, ML and DAGs provide a more efficient and open method for simulating complex conditions (Piccininni et al., n.d.).

Women from developing countries are not equally represented among AI-driven oral health datasets, which limits the development of appropriate and efficient healthcare solutions for a sizable portion of the population (Marko et al., 2025). This study aims to bridge that gap by investigating and predicting oral hyperpigmentation in pregnant and non-pregnant women aged 18 to 45, using DAG-supported ML models. This study contributes to more inclusive, data-driven maternal healthcare by developing predictive models tailored to the ethnically diverse population, aiming to improve patient counselling, improve early detection, and guide preventive measures in oral healthcare for females by addressing the relationship between age, occupation, ethnicity, and pregnancy status (Nyariro et al., 2023).

## Objectives

The objective of the study is to predict the association between oral hyperpigmentation and pregnancy utilizing multiple ML algorithms. This association will be validated in causal terms through DAGs for improving model interpretability and accuracy.

## Purpose

We hypothesized that ML models, supported by DAGs, can more meaningfully predict oral hyperpigmentation and identify key contributing factors such as age, occupation, ethnicity, and number of pregnancies.

## Methods

### Sample Description and Study Framework

In this cross-sectional study, 447 women within the ages of 18 and 45 were included from a public and private hospital through convenience sampling. The study was conducted from September 2021 to September 2024.

### Sample Size

Based on a probable population of 20,000, a 50% response distribution, a 96% confidence level, and a 5% margin of error, a sample size of 414 was calculated using the Raosoft sample size calculator (http://www.raosoft.com/samplesize.html). To further reduce the margin of error to 4.8%, we took a sample size of 447 students.

### Inclusion criteria

Pregnant and non-pregnant participants aged 18 to 45 years who were not suffering from any comorbid conditions were included in the study (Rezazadeh et al., 2014).

### Exclusion criteria

The exclusion criteria of the current study included factors such as smoking history (Tadakamadla et al., 2012), systemic diseases (Addison’s disease, melanoma), and medications known to cause hyperpigmentation (e.g., hydroxychloroquine) (Silva et al., 2022).

### DAGs Construction

DAGs depicted the relationship between oral hyperpigmentation and variables such as age, ethnicity, occupation, and number of pregnancies, as shown in Figure 1. DAG 1 shows the effect of ethnicity, age and number of pregnancies on oral hyperpigmentation, whereas DAG 2 shows the effect of occupation, age and ethnicity on oral hyperpigmentation. Mediating variables like occupation in DAG 2 link the primary exposure variable to the outcome variable. Demographic variables were treated as confounding variables, which were controlled for in ML models to prevent bias.

**Figure 1.**
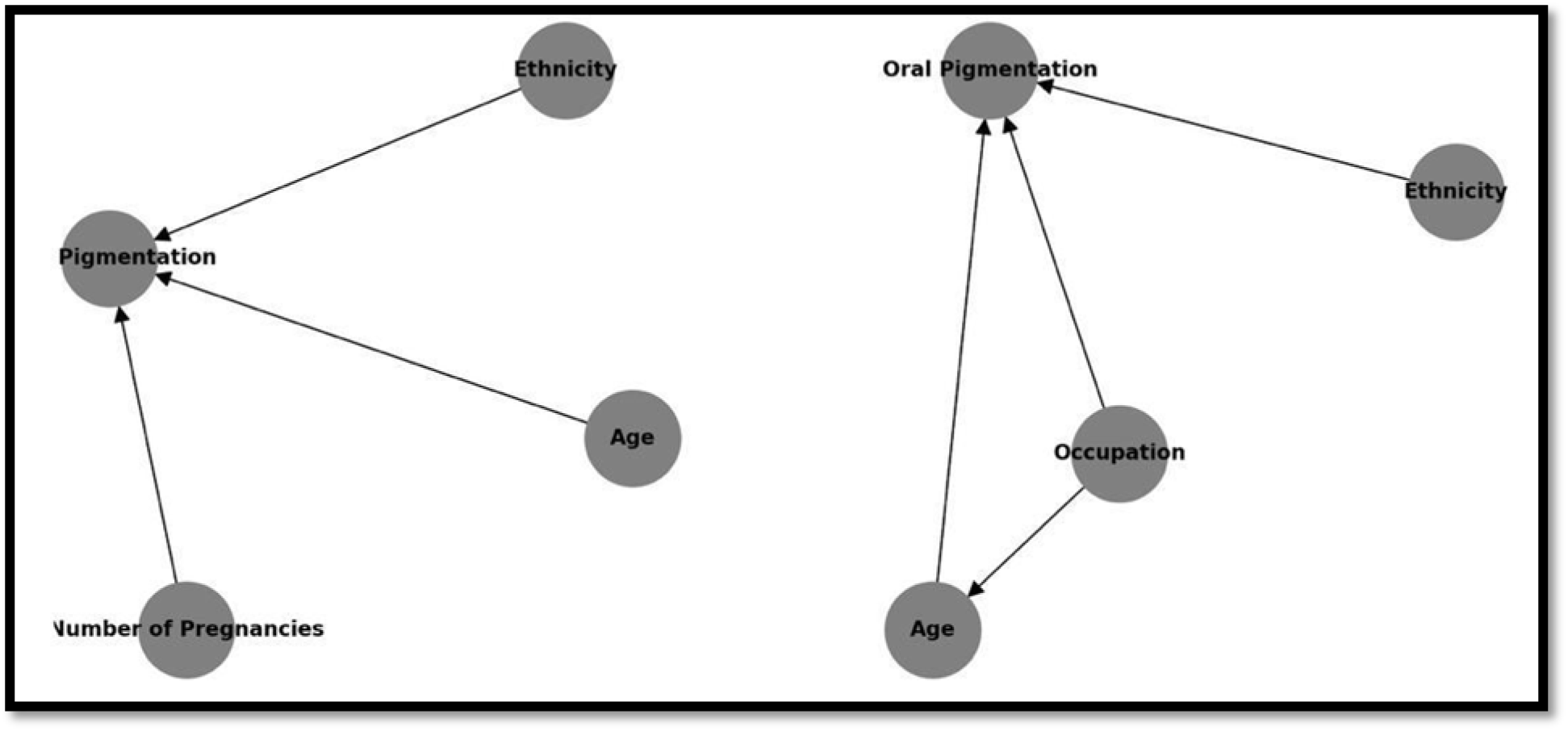
DAG model showing causal relationships influencing oral pigmentation in pregnancy. a. DAG 1 represents the effect of ethnicity, age, and number of pregnancies on oral hyperpigmentation. b. DAG 2 represents the effect of ethnicity, age, and occupation on oral hyperpigmentation.

### Data Collection

A standardized data collection form was designed to record both demographic details and clinical indicators of oral hyperpigmentation. Prior to the participants fulfilling the questionnaire, their written and informed consent was obtained. The factors recorded were age, ethnicity, occupation, number of pregnancies, presence or absence of oral hyperpigmentation, and specific hyperpigmentation across various oral sites like labial mucosa, soft palate, buccal mucosa, hard palate, floor of the mouth, and gingiva.

The Dummett Oral Hyperpigmentation Index (DOPI) was used to detect the presence and intensity of hyperpigmentation. The scoring pattern of DOPI (Muruppel et al., 2020) is as follows:

- No hyperpigmentation; indicated pink tissue, signifying the absence of clinical hyperpigmentation.
- Mild hyperpigmentation indicated light brown discoloration.
- Moderate hyperpigmentation indicated medium brown discoloration.
- Heavy hyperpigmentation indicated deep brown or blue-black discoloration.

Trained dental professionals with proper lighting conditions conducted clinical examinations to ensure accuracy and reduce inter-examiner variability. The inter-rater agreement was recorded as 90%. The hyperpigmentation scores were recorded on-site during the assessment. At least three of the five researchers evaluated each participant, and if there was a difference of opinion, the results were scrutinised to reach a unanimous decision. The final decision regarding unsettled differences was to be made by a fourth, senior-most examiner.

The variables of age and number of pregnancies were recorded as continuous variables, whereas ethnicity (balochi, kashmiri, pathan, punjabi, and sindhi) and occupation (student, housewife, and job) were taken as categorical variables, and oral hyperpigmentation as binary variables (present, absent). Oral hyperpigmentation of the gingiva, buccal mucosa, labial mucosa, hard palate, soft palate, and floor of the mouth were all assessed as ordinal variables (no hyperpigmentation, mild hyperpigmentation, moderate hyperpigmentation, and severe hyperpigmentation). To ensure data integrity and precise statistical analysis, both self-reported and clinically observed data were systematically collected.

### Statistical Analysis

Using Jamovi, Chi-square tests and Independent T-tests were applied to analyse the variations between categorical and continuous data, respectively (Kardys et al., 2013; Sundar & H., 2024). The results were shown with corresponding 95% confidence intervals where appropriate, and statistical significance was evaluated at p < 0.05. Three ML algorithms were employed: Gradient Boosting Machine (GBM), Logistic Regression (LR), and Random Forest (RF) to model the binary outcome variable—presence or absence of oral hyperpigmentation. These models were utilized because they have proven abilities to interpret high-dimensional data and detect complex associations between variables. Preprocessing methods, such as one-hot encoding of categorical data and normalization of continuous variables, were done for the implementation model. A training/test ratio of 80:20 was used on the dataset. GridSearch cross-validation (CV) was used for hyperparameter tuning. The precision, recall, accuracy, the F1 score, and the extent under the ROC curve (AUC-ROC) were among the performance criteria for evaluation.

Correlation analyses were done after stratification of the data to analyze the link between continuous variables. Correlation matrices were generated and shown by heatmaps. Figure 2 shows heatmaps in cool-warm colour schemes to highlight positive and negative correlations.

**Figure 2.**
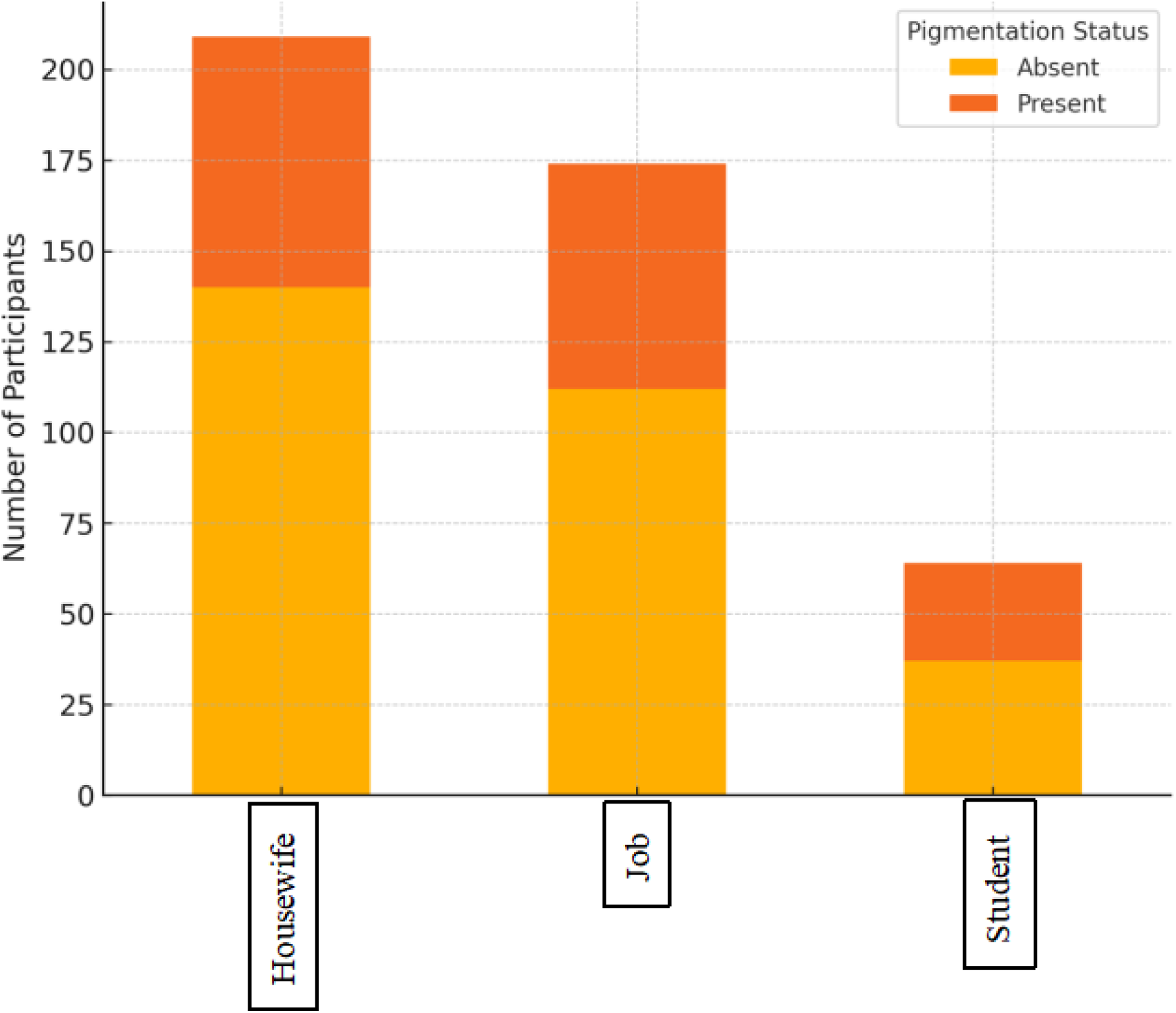
Bar graph showing pigmentation status between three study groups based on occupation.

To ensure data integrity, missing values were handled using appropriate imputation techniques. If more than 10% of the participants’ data were missing, that entry was excluded from analysis to prevent bias.

### Statistical Software Used

The analyses utilized Python (version 3.12) libraries, such as *Scikit-Learn,* like *Pandas* for analyzing large datasets, *Seaborn* for statistical data visualizations and correlations, and *NumPy* for numerical calculations (Wornow et al., 2023).

### Ethical Approval

The university’s Research Ethics Committee provided ethical authorisation for the study. In order to minimise selection bias, participants who matched the inclusion criteria and provided written informed consent were included in the study. On reasonable request, the corresponding author will hand over the code and dataset utilised during the research.

## Results

### Descriptive statistics

According to the data shown in Table 1, 447 participants were selected for the study, with their numbers of pregnancies ranging between 0 and 5. Among them, 289 (64.7%) had no oral hyperpigmentation, while 158 (35.3%) had oral hyperpigmentation. The distribution of hyperpigmentation showed no statistically significant association with the number of pregnancies (p = 0.736), although a slight increase in hyperpigmentation was observed among those with multiple pregnancies.

**Table 1.** Descriptive Statistics.

| Number of Pregnancies |  |  |  |  |  |  |  |  |
| --- | --- | --- | --- | --- | --- | --- | --- | --- |
|  | 0<br>N (%) | 1<br>N (%) | 2<br>N (%) | 3<br>N (%) | 4<br>N (%) | 5<br>N (%) | Total<br>N (%) | P-value |
| **Oral Hyperpigmentation** |  |  |  |  |  |  |  |  |
| Absent | 78<br>(27%) | 54<br>(18.7%) | 62<br>(21.5%) | 58<br>(20.1%) | 26<br>(9%) | 11<br>(3.8%) | 289<br>(100%) | 0.736 |
| Present | 45<br>(28.5%) | 21<br>(13.3%) | 39<br>(24.7%) | 34<br>(21.5%) | 12<br>(7.6%) | 7<br>(4.4%) | 158<br>(100%) |  |
| Total | 123<br>(27.5%) | 75<br>(16.8%) | 101<br>(22.6%) | 92<br>(20.6%) | 38<br>(8.5%) | 18<br>(4.0%) | 447<br>(100%) |  |
| **Labial Mucosa** |  |  |  |  |  |  |  |  |
| No Hyperpigmentation | 115<br>(29.6%) | 68<br>(17.5%) | 85<br>(21.9%) | 78<br>(20.1%) | 28<br>(7.2%) | 14<br>(3.6%) | 388<br>(100%) | 0.003 |
| Mild Hyperpigmentation | 8<br>(14.3%) | 7<br>(12.5%) | 16<br>(28.6%) | 13<br>(23.2%) | 9<br>(16.1%) | 3<br>(5.4%) | 56<br>(100%) |  |
| Moderate Hyperpigmentation | 0<br>(0%) | 0<br>(0%) | 0<br>(0%) | 1<br>(50%) | 0<br>(0%) | 1<br>(50%) | 2<br>(100%) |  |
| Heavy Hyperpigmentation | 0<br>(0%) | 0<br>(0%) | 0<br>(0%) | 0<br>(0%) | 1<br>(100%) | 0<br>(0%) | 1<br>(100%) |  |
| Total | 123<br>(27.5%) | 75<br>(16.8%) | 101<br>(22.6%) | 92<br>(20.6%) | 38<br>(8.5%) | 18<br>(4.0%) | 447<br>(100%) |  |
| **Hard Palate** |  |  |  |  |  |  |  |  |
| No Hyperpigmentation | 123<br>(28%) | 73<br>(16.6%) | 101<br>(23.0%) | 88<br>(20.0%) | 38<br>(8.6%) | 17<br>(3.9%) | 440<br>(100%) | 0.018 |
| Mild Hyperpigmentation | 0<br>(0%) | 2<br>(28.6%) | 0<br>(0%) | 4<br>(57.1%) | 0<br>(0%) | 1<br>(14.3%) | 7<br>(100%) |  |
| Total | 123<br>(27.5%) | 75<br>(16.8%) | 101<br>(22.6%) | 92<br>(20.6%) | 38<br>(8.5%) | 18<br>(4.0%) | 447<br>(100%) |  |
| **Floor of The Mouth** |  |  |  |  |  |  |  |  |
| No Hyperpigmentation | 123<br>(27.7%) | 75<br>(16.9%) | 101<br>(22.7%) | 90<br>(20.3%) | 38<br>(8.6%) | 17<br>(3.8%) | 444<br>(100%) | 0.044 |
| Mild Hyperpigmentation | 0<br>(0%) | 0<br>(0%) | 0<br>(0%) | 2<br>(66.7%) | 0<br>(0%) | 1<br>(33.3%) | 3<br>(100%) |  |
| Total | 123<br>(27.5%) | 75<br>(16.8%) | 101<br>(22.6%) | 92<br>(20.6%) | 38<br>(8.5%) | 18<br>(4.0%) | 447<br>(100%) |  |

Labial mucosa hyperpigmentation demonstrated a statistically significant association with the number of pregnancies (p = 0.003). Most participants (388, 86.8%) had no labial hyperpigmentation, 56 had light hyperpigmentation (12.5%), and only 3 (0.7%) had moderate to severe hyperpigmentation.

Additionally, there were significant differences in hard palate hyperpigmentation between pregnancy groups (p = 0.018). Only 7 people (1.6%) had insignificant hyperpigmentation on the hard palate, mostly in those with one to three pregnancies, while 440 people (98.4%) had no visible hyperpigmentation at all.

Hyperpigmentation on the floor of the mouth was less frequent but still statistically significant (p-value= 0.044). Mild hyperpigmentation was reported in 3 participants (0.7%), all of whom had experienced 3 or more pregnancies.

As shown in Table 1, the results showed a p-value of 0.736, which indicated an insignificant association between the oral hyperpigmentation and the number of pregnancies. Hyperpigmentation on the labial mucosa and hard palate, having p-values of 0.003 and 0.018, respectively, showed significance with the number of pregnancies.

According to Figure 2, oral hyperpigmentation was more commonly observed among housewives and individuals with jobs compared to students. The higher hyperpigmentation levels in housewives and employed participants may be attributed to greater exposure to environmental or lifestyle-related factors.

### Correlation Analysis

The Pearson coefficients for the correlation (R) between the main factors affecting oral hyperpigmentation are shown in Table 2. Age and the number of pregnancies have been found to be slightly positively correlated (R = 0.60). There was a moderate correlation between gingival hyperpigmentation and buccal mucosa (R = 0.38) but a significant correlation between gingival hyperpigmentation and overall oral hyperpigmentation (R = 0.86). Figure 3 shows a correlation heatmap illustrating relationships between age, number of pregnancies, and oral hyperpigmentation. Values represent Pearson correlation coefficients, color-coded on a cool-warm scale, where red indicates stronger positive associations and blue indicates weaker or negative ones.

**Figure 3.**
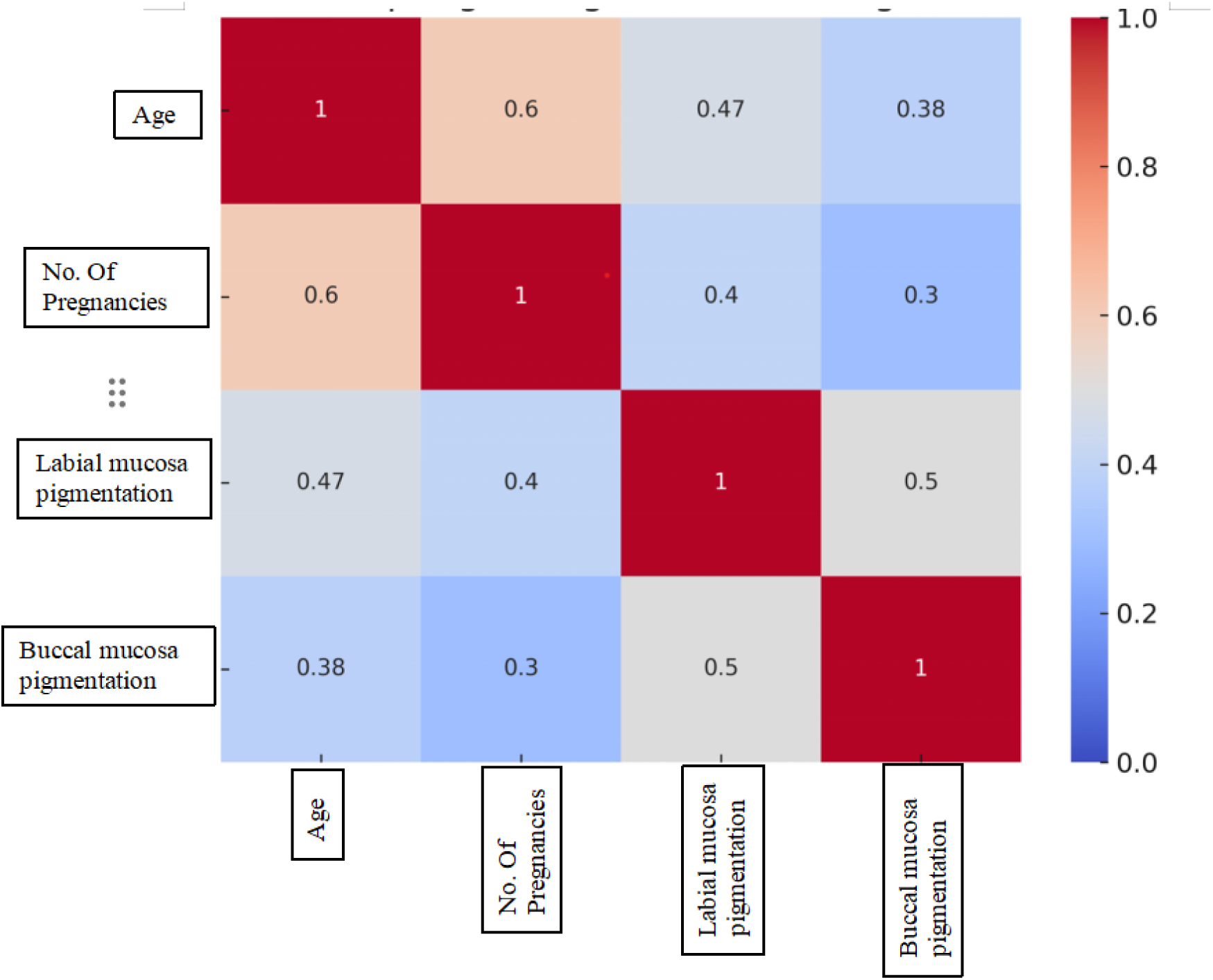
Correlation Heatmap between age, number of pregnancies, and hyperpigmentation on labial and buccal mucosa.

**Table 2.** Variable Associations Analysis.

| Correlation of Key Variables | Correlation Coefficient (R) |
| --- | --- |
| Age and Number of Pregnancies | 0.6 |
| Total Hyperpigmentation and Gingiva | 0.86 |
| Total Hyperpigmentation and Labial Mucosa | 0.47 |
| Total Hyperpigmentation and Buccal Mucosa | 0.38 |

### ML Model Metrics

Three ML models were applied to predict oral hyperpigmentation according to age, occupation, ethnicity and pregnancy status. All three ML models demonstrated excellent predictive power. LR provided the best performance with an accuracy of 96.67%; RF and GBM came a close second (both 95.56%). For all three models, precision, recall and F1 scores were similarly high, with LR performing particularly well, as shown in Table 3.

**Table 3.** Machine Learning Model Metrics.

| Model | Accuracy | Precision | Recall | F1 Score | AUC-ROC |
| --- | --- | --- | --- | --- | --- |
| Random Forest | 0.9556 | 0.9667 | 0.9063 | 0.9355 | 0.9445 |
| Gradient Boosting Machine | 0.9556 | 0.9667 | 0.9063 | 0.9355 | 0.9445 |
| Logistic Regression | 0.9667 | 0.9677 | 0.9375 | 0.9524 | 0.9601 |

### ROC-AUC Graphs

ROC-AUC scores indicated that LR had the strongest discriminatory ability with an AUC of 0.9601, compared to 0.9445 for RF and GBM. The ROC curves (Figure 4) show that all models performed well, though LR demonstrated marginally better performance at various thresholds.

**Figure 4.**
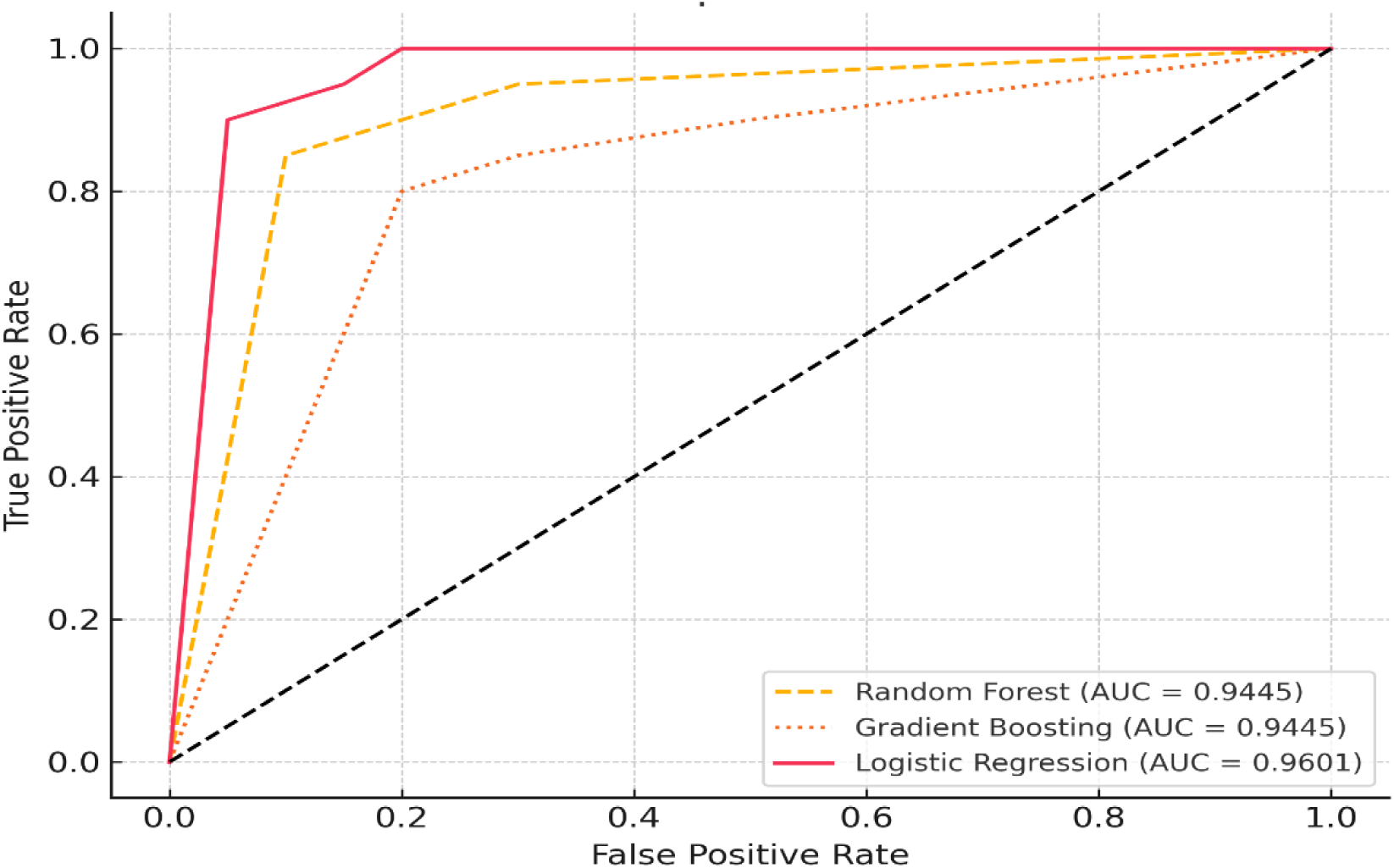
ROC curve showing model performance for pigmentation prediction.

## Discussion

The current study reported an insignificant association between the total presence of oral hyperpigmentation and the number of pregnancies. However, localized hyperpigmentation at the hard palate, floor of the mouth, and labial mucosa showed significant correlation with pregnancy. These findings support the hormonal theories, especially those that link melanin synthesis to higher levels of progesterone and estrogen during pregnancy (Snell & Bischitz, 1960).

### Age and Ethnicity

The number of pregnancies was moderately correlated with age and ethnicity (Rezazadeh et al., 2014). These findings and the model calculations are in accordance with findings from other studies (Koppolu et al., 2024). For example, in South Africa, a hyperpigmentation prevalence of 42% among darker skin tone individuals is consistent with our findings that some ethnic groups in developing countries are more likely to have oral hyperpigmentation (Masilana et al., 2017).

### Environmental and Occupational Factors

Compared to students, housewives and working females showed higher levels of hyperpigmentation, which showed occupation as a possible contributing factor, maybe as a result of increased exposure to environmental pollutants or stressors associated with their lifestyles. Based on the literature, some occupational categories are particularly affected by environmental factors such as smoking (Moravej-Salehi et al., 2015) and sun exposure (Lim et al., 2025).

### Machine Learning and DAGs

All three ML models demonstrated excellent predictive power. However, the LR model with an accuracy rate of 96.67% was particularly effective, whereas GBM and RF with an AUC of 0.9601 slightly underperformed. The underperformance of RF and GBM may be due to overfitting, inadequate data complexity, or not having enough training data, which are factors that are consistent with previous studies on small-sample ML applications in health domains (Han et al., 2021). In contrast, LR benefitted from its interpretability and ability to generalize well on relatively smaller datasets (Nusinovici et al., 2020).

### Under-representation of females from developing countries in AI models

In comparison with global models based on datasets that mainly focus on Western populations, this study confronts the significant gap in representation of females from developing countries in AI-driven oral health research via a prediction tool that considered the genetic and socio-environmental characteristics. Studies from South Asian ethnicities, such as women from Bangladesh, Nepal, or India, demonstrated high hyperpigmentation variation associated with skin tone and reproductive status, signifying that it might be beneficial (Rijal et al., 2021; Ponnaiyan et al., 2014). To validate and adjust the model for wider South Asian use, comparative research needs to be encouraged (Rotbeh et al., 2022).

### Clinical and Public Health Implications and Recommendations

Real-time risk assessments can be obtained by integrating ML into electronic health records (EHRs), which can help direct interventions and referrals, especially for expectant mothers (Lipschuetz et al., 2024). Early diagnosis of pregnancy-related oral hyperpigmentation, such as on the labial mucosa and hard palate, may improve treatment compliance and minimise anxiety. While the LR method helps in identifying high-risk women and expedites education and referrals, culturally tailored explanations and community programs can improve involvement.

### Strengths and Limitations

The limitations of the study included its cross-sectional design and dependency on self-reported demographic data, which could result in reporting bias or recall error. Likewise, environmental factors, including smoking status as well as UV exposure, were not recorded, which would have led to the exclusion of significant hyperpigmentation contributors.

The strengths of the study included the utilization of trained dentists for hyperpigmentation assessment, detailed information gathered using predefined clinical indicators, and an innovative blended utilization of ML and DAGs to further improve understanding and prediction effectiveness. In comparison to prior studies, this study resolves a number of significant limitations. We addressed the more generic age sample used in past research by focusing on women within the ages of 18 and 45, allowing us to evaluate pregnancy-related oral hyperpigmentation in a demographically relevant cohort. Overall, this study highlights important topics for further research while providing a more focused and methodologically rigorous examination of oral hyperpigmentation in women from developing countries. To improve reproducibility, the study applied explicit methods for analysis incorporating openly accessible software (Jamovi, Python-based ML libraries) and standard machine learning methods (normalization, encoding, GridSearch CV). Prospective research might look into open-access dissemination of data and software availability that would allow worldwide comparison and validation by other researchers.

## Conclusions

This study shows how ML algorithms can potentially be applied for accurately assessing oral hyperpigmentation in women within the age range of 18 and 45 when reinforced by DAGs. The results emphasize the importance of targeted methods of diagnosis, primarily in areas with limited resources and highlight the complicated associations among tailored oral hyperpigmentation and pregnancy, ethnic background, age, and occupation. To improve generalizability, additional studies should include psychological and environmental factors, contrasts to comparable groups, and broader geographical aspects.

## Author Acknowledgements

Arooba Malik and Hifza Noor contributed to the original draft, writing, methodology, data collection, conceptualization, references, review and editing. Faisal Shafiq and Mahnoor Fatima contributed to the review and editing. Rameesa Aymen and Momina Zahra contributed to data collection. The study was not funded by any external organization.

## Data Availability

All data produced in the present study are available upon reasonable request.

## References

Abati, S., Sandri, G. F., Finotello, L., & Polizzi, E. (2024). Differential Diagnosis of Pigmented Lesions in the Oral Mucosa: A Clinical Based Overview and Narrative Review. Cancers, 16(13), 2487. 10.3390/cancers16132487

Agarwal, M. C., Agarwal, S., Madan, E., & Madan, R. (2015). GINGIVAL HYPERPIGMENTATION REVISITED. https://www.researchgate.net/publication/361585336

Ciçek, Y., & Ertaş, U. (2003). The normal and pathological hyperpigmentation of oral mucous membrane: a review. The Journal of Contemporary Dental Practice, 4(3), 76–86.

Pecci-Lloret, M. P., Linares-Pérez, C., Pecci-Lloret, M. R., Rodríguez-Lozano, F. J., & Oñate-Sánchez, R. E. (2024). Oral Manifestations in Pregnant Women: A Systematic Review. Journal of Clinical Medicine, 13(3). 10.3390/jcm13030707

Rotbeh, A., Kazeminia, M., Kalantari, M., & Rajati, F. (2022). Global prevalence of oral hyperpigmentation and its related factors: a systematic review and meta-analysis. *Journal of Stomatology*, Oral and Maxillofacial Surgery, 123(5), e411–e424. 10.1016/j.jormas.2022.01.009

Gulati, N., Dutt, P., Gupta, N., & Tyagi, P. (n.d.). GINGIVAL HYPERPIGMENTATION: REVISITED. In Journal of Advanced Medical and Dental Sciences Research | Vol (Vol. 4).

Masilana, A., Khammissa, R. A. G., Lemmer, J., & Feller, L. (2017). Physiological oral melanin hyperpigmentation in a South African sample: A clinical study. Journal of Investigative and Clinical Dentistry, 8(4). 10.1111/jicd.12258

Ponnaiyan, D., Jegadeesan, V., Perumal, G., & Anusha, A. (2014). Correlating skin color with gingival hyperpigmentation patterns in South Indians - a cross sectional study. Oral Health and Dental Management, 13(1), 132–136.

Feller, L., Masilana, A., Khammissa, R. A., Altini, M., Jadwat, Y., & Lemmer, J. (2014). Melanin: the biophysiology of oral melanocytes and physiological oral hyperpigmentation. Head & Face Medicine, 10(1), 8. 10.1186/1746-160X-10-8

Soulafa Almazrooa, Amna Arooj, Sobia Hassan, & Amber Kiyani. (2025). JOHOE_Volume 14_Issue 1_Pages 1-5. Journal of Oral Health and Oral Epidemiology, 14, 1–5. 10.34172/johoe.2403.1627

Shahzad, A., Kiyani, A., & Paiker, S. (2018). Prevalence of Oral Anomalies and Pathologies in the Pakistani Population – A Cross Sectional Study. Journal of The Pakistan Dental Association, 27(1), 13–17. 10.25301/JPDA.271.13

Motosko, C. C., Bieber, A. K., Pomeranz, M. K., Stein, J. A., & Martires, K. J. (2017). Physiologic changes of pregnancy: A review of the literature. International Journal of Women’s Dermatology, 3(4), 219–224. 10.1016/j.ijwd.2017.09.003

Sattar, U., Malik, U., Shireen, Z., Faiz Ahmad Khan, S., Health CenterQadirabad, R., & Ghazi Khan, D. (2017). Oral health challenges in pregnant women in Pakistan: A research analysis. International Journal of Advanced Biotechnology and Research (IJBR), 8, 1724–1730. http://www.bipublication.com

Dubuc, A., Zitouni, A., Thomas, C., Kémoun, P., Cousty, S., Monsarrat, P., & Laurencin, S. (2022). Improvement of Mucosal Lesion Diagnosis with Machine Learning Based on Medical and Semiological Data: An Observational Study. Journal of Clinical Medicine, 11(21), 6596. 10.3390/jcm11216596

Piccininni, M., Konigorski, S., Rohmann, J. L., & Kurth, T. (n.d.). *Title:* Directed Acyclic Graphs and causal thinking in clinical risk prediction modeling.

Marko, J. G. O., Neagu, C. D., & Anand, P. B. (2025). Examining inclusivity: the use of AI and diverse populations in health and social care: a systematic review. BMC Medical Informatics and Decision Making, 25(1), 57. 10.1186/s12911-025-02884-1

Nyariro, M., Emami, E., Caidor, P., & Abbasgholizadeh Rahimi, S. (2023). Integrating equity, diversity and inclusion throughout the lifecycle of AI within healthcare: a scoping review protocol. BMJ Open, 13(9), e072069. 10.1136/bmjopen-2023-072069

Rezazadeh, F., Falsafi, N., Sarraf, Z., & Shahbazi, M. (2014). Oral Mucosal Disorders in Pregnant versus Non-Pregnant Women. Dentistry Journal, 2(4), 134–141. 10.3390/dj2040134

Tadakamadla, J., Kumar, S., Nagori, A., Tibdewal, H., Duraiswamy, P., & Kulkarni, S. (2012). Effect of smoking on oral hyperpigmentation and its relationship with periodontal status. Dental Research Journal, 9(Suppl 1), S112–4.

Silva, P. U. J., Oliveira, M. B., Vieira, W., Cardoso, S. V., Blumenberg, C., Franco, A., Siqueira, W. L., & Paranhos, L. R. (2022). Oral hyperpigmentation as an adverse effect of chloroquine and hydroxychloroquine use. Medicine, 101(11). 10.1097/MD.0000000000029044

Kardys, I., Hoeks, S., van Domburg, R., Lenzen, M., & Boersma, E. (2013). Tools and techniques - Statistics: Analysis of continuous data using the t-test and ANOVA. In EuroIntervention (Vol. 9, Issue 6, pp. 765–767). 10.4244/EIJV9I6A123

S., V., A.S, H., & S. Sundar, J. (2024). CHI-SQUARE TESTS: A QUICK GUIDE FOR HEALTH RESEARCHERS. International Journal of Advanced Research, 12(10), 1214–1222. 10.21474/IJAR01/19746

Wornow, M., Gyang Ross, E., Callahan, A., & Shah, N. H. (2023). APLUS: A Python library for usefulness simulations of machine learning models in healthcare. Journal of Biomedical Informatics, 139, 104319. 10.1016/j.jbi.2023.104319

Muruppel, A. M., Pai, B. S. J., Bhat, S., Parker, S., & Lynch, E. (2020). Laser-Assisted Dehyperpigmentation—An Introspection of the Science, Techniques, and Perceptions. Dentistry Journal, 8(3), 88. 10.3390/dj8030088

Snell, R. S., & Bischitz, P. G. (1960). The Effect of Large Doses of Estrogen and Estrogen and Progesterone on Melanin Hyperpigmentation**From the Department of Anatomy, Medical School King’s College, Newcastle upon Tyne, University of Durham, England. Journal of Investigative Dermatology, 35(2), 73–82. 10.1038/jid.1960.87

Rezazadeh, F., Falsafi, N., Sarraf, Z., & Shahbazi, M. (2014). Oral Mucosal Disorders in Pregnant *versus* Non-Pregnant Women. Dentistry Journal, 2(4), 134–141. 10.3390/dj2040134

Koppolu, P., Almutairi, H., Yousef, S. al, Ansary, N., Noushad, M., Vishal, M. B., Swapna, L. A., Alsuwayyigh, N., Albalawi, M., Shrivastava, D., & Srivastava, K. C. (2024). Relationship of skin complexion with gingival tissue color and hyperhyperpigmentation. A multi-ethnic comparative study. BMC Oral Health, 24(1), 451. 10.1186/s12903-024-04189-7

Moravej-Salehi, E., Moravej-Salehi, E., & Hajifattahi, F. (2015). Relationship of Gingival Hyperpigmentation with Passive Smoking in Women. Tanaffos, 14(2), 107–114.

Lim, H. W., Piquero-Casals, J., Schalka, S., Leone, G., Trullàs, C., Brown, A., Foyaca, M., Gilaberte, Y., Krutmann, J., & Passeron, T. (2025). Photoprotection in pregnancy: addressing safety concerns and optimizing skin health. Frontiers in Medicine, 12. 10.3389/fmed.2025.1563369

Han, S., Williamson, B. D., & Fong, Y. (2021). Improving random forest predictions in small datasets from two-phase sampling designs. BMC Medical Informatics and Decision Making, 21(1), 322. 10.1186/s12911-021-01688-3

Nusinovici, S., Tham, Y. C., Chak Yan, M. Y., Wei Ting, D. S., Li, J., Sabanayagam, C., Wong, T. Y., & Cheng, C.-Y. (2020). Logistic regression was as good as machine learning for predicting major chronic diseases. Journal of Clinical Epidemiology, 122, 56–69. 10.1016/j.jclinepi.2020.03.002

Rijal, A. H., Dhami, B., Pandey, N., & Aryal, D. (2021). Prevalence of Gingival Hyperpigmentation and its Association with Gingival Biotype and Skin Colour. Journal of Nepalese Society of Periodontology and Oral Implantology, 5(1), 19–25. 10.3126/jnspoi.v5i1.38178

Lipschuetz, M., Guedalia, J., Cohen, S. M., Unger, R., Yagel, S., & Sompolinsky, Y. (2024). Machine Learning in Electronic Health Records: Identifying High-Risk Obstetric Patients Pre and During Labor. 10.3233/SHTI240096

